# Passive surveillance of human-biting ticks correlates with town-level disease rates in Massachusetts

**DOI:** 10.1101/2022.05.01.22274432

**Authors:** Nathalie Lavoie, Guang Xu, Catherine Brown, Michel Ledizet, Stephen M. Rich

**Affiliations:** L2 Diagnostics, LLC. New Haven, Connecticut; Laboratory of Medical Zoology, Department of Microbiology, University of Massachusetts-Amherst, Amherst, Massachusetts; Massachusetts Department of Public Health, Boston, Massachusetts

**Keywords:** Ixodes scapularis, Disease risk, Lyme disease, Babesiosis, Anaplasmosis, Borrelia miyamotoi, Passive tick surveillance

## Abstract

We assessed the temporal and spatial distribution of *Borrelia burgdorferi, Borrelia miyamotoi, Babesia microti*, and *Anaplasma phagocytophilum* among human-biting *Ixodes scapularis* ticks in Massachusetts using ticks submitted to the TickReport pathogen passive surveillance program. From January 2015 to December 2017, *Ixodes scapularis* was the most frequently submitted tick species (n=7462). *B. burgdorferi* prevalence increased in ticks during the study period in adults and nymphs (37.1-39.1% in adults, 19.0%-23.9% in nymphs). The proportion of *B. microti* infected ticks increased from 5.7% to 8.1% in adult ticks but remained constant in nymphs (5.4-5.6%). Stable or decreasing annual prevalence of *B. miyamotoi* (2.2 – 2.2% in adults, 1.0-1.9% in nymphs) and *A*.*phagocytophilum* (7.6-7.2% in adults, 5.0-4.0% in nymphs) were detected. Coinfections were observed and included all pathogen combinations.

Ticks were submitted year-round and had stable infection rates. The temporal pattern of *B. burgdorferi-* positive nymphs aligned with reported cases of Lyme disease, as did positive *B. microti* nymphs and babesiosis. A similar situation is seen with *B. miyamotoi* with an insignificant fall peak in cases. Anaplasmosis demonstrated a significant bimodal distribution with reported cases peaking in the spring and fall. This pattern is similar to that of *A. phagocytophilum-*infected adult ticks.

*B. microti* infected nymphs were significantly predictive of town-level babesiosis incidence and *A. phagocytophilum* infected adults were significantly predictive of town-level anaplasmosis incidence in a spatially adjusted negative binomial model. Unlike field collection studies, the high number of ticks submitted provides a high-resolution picture of pathogen prevalence and provides data relevant to human health at the town level. Through temporal and geographic analyses we demonstrate concordance between our passive surveillance tick pathogen data and state reports of tickborne disease.

## Introduction

*Ixodes scapularis* is the primary vector for a variety of pathogens endemic to the northeast United States causing a heavy burden of human illness in the northeast United States. Chief among these illnesses is Lyme disease, caused by *Borrelia burgdorferi* sensu stricto which accounts for an estimated >300,000 cases in the United States (Nelson et al.). *B. microti*, though asymptomatic in most immunocompetent hosts, poses a substantial threat to the blood supply in its endemic region. Infection in immunocompromised hosts and the elderly results in clinical manifestations similar to malaria and confers serious risk of hospitalization and possible fatality (Vannier et al., 2015). Human granulocytic anaplasmosis was documented in Massachusetts in 1995 and nationally, reports have increased from 348 cases in 2000, to 4,151 cases in 2016 (Telford et al., 1995, 2020). The emerging pathogen *B. miyamotoi* is not yet a nationally reportable disease although it was made notifiable in Massachusetts in 2014. The transmission rate of *B. miyamotoi* to humans by infected ticks is reportedly greater than the corresponding rate of transmission of *B. burgdorferi*, indicating the potential for further emergence and disease burden (Sarksyan et al., 2015). Reports of suspect cases in Massachusetts have increased from 12 in 2014 to 191 in 2018 (MASDOH, unpublished). Awareness of the pathogen and associated clinical presentations remain limited which may contribute to under-reporting.

TickReport is a tick testing service based at the Laboratory of Medical Zoology at UMass Amherst. Subscribers to TickReport submit their ticks and receive a report on three critical factors of risk of human diseases. The first risk factor reported is the tick species. CDC advises that people identify the species of tick because not all ticks are associated with particular pathogens (CDC, 2019). The second risk factor evaluated is the degree of feeding. Pathogen transmission only occurs after ticks have fed for some time, though the precise timing varies for different pathogen species (Eisen, 2018). The third risk factor determined by TickReport is the infection status for each of the most common pathogens common to the human-biting tick species. While CDC does not recommend tick testing, many tick-bite victims find it valuable to receive this information to aid in risk assessment. The aggregated TickReport data comprise a valuable passive surveillance database for measuring the exposure risk to human populations by direct accounting of human-biting ticks.

Here we have utilized these passive surveillance data to explore the geographic and temporal trends in *Borrelia burgdorferi, Borrelia miyamotoi, Babesia microti*, and *Anaplasma phagocytophilum* infection in ticks in Massachusetts from 2015 to 2017. Historically, research on these pathogens in their vector, *Ixodes scapularis*, has relied on entomological sampling of ticks in the wild by flagging or dragging. These methods are subject to bias and are generally limited in their geographic and temporal coverage. In addition, ticks collected through these methods may not have a high level of interaction with human hosts, reducing the ability to draw direct inference on public health risk. In contrast, greater than 95% of ticks analyzed in this project are collected from humans. We demonstrate the utility of these data by aligning information on tick with cases of tick-borne diseases reported to the Massachusetts Department of Public Health.

TickReport passive surveillance presents an invaluable resource for interrogating important questions about health risks associated with *Ixodes scapularis* tick bites. Here we report on that passive surveillance in Massachusetts, where we compared prevalence of infection among human-biting ticks with *B. burgdorferi, B. miyamotoi, A. phagocytophilum*, and *B. microti*. Our data demonstrate increases in frequency of two of these pathogens over the study period. Data from TickReport offer an improved surveillance methodology by including only ticks known to have interacted with human hosts, with known engorgement levels, and at a high spatial resolution across the state of Massachusetts. We demonstrate geographic and temporal concordance between pathogens in ticks and reported tickborne diseases. The pattern seen with anaplasmosis provides critical information about the importance of adult ticks as vectors for transmission. We demonstrate that passive surveillance of ticks provides data to confirm and complement clinical case report information.

## Material and Methods

### Tick Collection via Submission

Methods of tick collection, identification, and PCR have been described in detail elsewhere [Xu et. al 2016]. Ticks were collected during January 2015 through December 2017 through the tick identification and pathogen service hosted at University of Massachusetts (www.umass.edu/tick). Metadata attached to the tick specimen included date of tick collection and the town where the user suspected the tick bite to have occurred. When this location was not given, home address was used to geotag ticks. We received tick specimens through postal mail at the University of Massachusetts Amherst closed in small plastic vials or zipper-locking bags. When mapping percent positive ticks per town, only towns that submitted greater than five ticks are shown. All towns with less than 5 ticks are shown in grey.

### Tick Identification and PCR

Tick species identification was based on published identification keys (Keirans et al., 1996) Keirans and Clifford 1978, Keirans and Litwak 1989). Ticks were categorized by developmental stage (larva, nymph, or adult) and engorgement levels (un-engorged, partially fed, and engorged). Differentiation of *I. scapularis* and *I. pacificus* was performed by a species-specific TaqMan PCR assay (Xu et al. 2019).

### Case Data

Municipality-level reported case data were obtained from the Massachusetts Department of Health and included data on cases of Lyme disease from 2015, and cases of anaplasmosis and babesiosis from 2015-2017. Case counts at the municipal level were aggregated over the three-year research period and statewide data were provided by month. To protect patients’ privacy, case counts in municipalities with between 1-4 reported cases per year were blinded. In these instances, a random value between 1 and 4 was assigned to each town. Data aggregated here met the confirmed or probable definition for each pathogen. These definitions can be found in full at the CDC. In short, a case requires laboratory evidence of infection by a test included in the standardized case definition and which varies by disease organism, and a report of a clinically compatible illness in the patient. Cases are assigned to a geography based on their official address and not based on their location of exposure to the tick-borne pathogen. Cases are also assigned temporally by the month of their symptom onset. Data on Lyme disease are limited to 2015 as active case report follow-up was discontinued. Therefore, no cases from 2016-2017 can be classified as confirmed or probable.

### Spatial Data and Geostatistics

Statistical and spatial analysis were performed using R software version 3.4.2 and the graphical user interface RStudio version 1.1.383 (R Core Team, 2013). Shapefiles for the town boundaries for the state of Massachusetts were obtained from MassGIS (MassGIS 2014). The use of the term “town” in this paper references a municipality as defined by MASSGIS. This data layer contains US Census Town Population from 2010 which was used as the population denominator for all three-year incidence calculations. All mapping was performed in R with the packages: rgdal (Bivand et al., 2017), sp (Pebesma & Bivand, 2005), geoR (Ribeiro & Diggle, 2016), spdep (Bivand & Wong, 2018), and maptools (Bivand & Lewin-Koh, 2017).

### Regression Model Construction

Regression models were used to investigate the relationship between reported cases of babesiosis and anaplasmosis per town and the presence of the respective pathogens in the tick vector. The R package MASS and function “glm.nb” was used to generate the negative binomial model (Venables & Ripley, 2002). Negative binomial model was used to account for overdispersion in a Poisson count approach. The goal of these models was to correlate infected ticks (input) at the town-level with the reported case count (output). This model also required variables to adjust for population; a log-offset of the 2010 Census population was used. To improve the models, a spatial element was incorporated.

### Spatial Element

It is assumed that both tick counts and case reports closer to each other are more similar than those further away. To quantify this spatial autocorrelation spatially-lagged variables were generated that determined that average of each dependent variable in the neighboring towns. The term “spatial lag” refers to an average of all towns that touch the border of each given town.

### Final Model Selection

The best fit model was selected by AIC. Akaike information criterion (AIC), a goodness of fit test based on the likelihood function of the model which incorporates a penalty for the inclusion of additional parameters. Only statistically significant variables were included in the final model, and log-likelihood test against null and step removal were used to confirm the importance of each variable.

### Coinfection Statistics

Confidence intervals were calculated by the procedure given by Clopper and Pearson (1934) using “binom.test” in the R software. Pearson’s Chi-square test for count data was performed in R using “chisq.test” in the stats package to determine the statistical independence of tick infection and coinfection. The test outcome indicates if significantly more (or less) coinfected ticks are observed than expected under statistical independence.

*Additional details on the geostatistical analysis can be found in the* ***Supplemental Methods***.

## Results

### Passive surveillance facilitates cost-effective tick collection and frequently captures singly and co-infected ticks of all stages

A total of 8,787 ticks were submitted with a reported location in Massachusetts (**Table 1**). Twelve species were identified in Massachusetts. The majority of ticks were *Ixodes scapularis* (85%), followed by *Demacentor variabilis* (12%) and *Amblyomma americanum* (1%). *Ixodes scapularis* were submitted from 335 out of the 351 total Massachusetts municipalities (95.4% coverage). The total number of tick submissions per town for adults and nymphs is shown in **Figure 1**. For the sake of visualization, a cut-off of five ticks per town was required for town inclusion in maps. This allowed 70% coverage with 234 towns submitting greater than 5 ticks during the study period. Samples were submitted in every month of the three-year complete study period except for February 2015. In 96.3% of submissions, Massachusetts was the home state of the individual and 72% of all submissions reported tick bites occurred within the same town as their home address. Adults (95.2%), nymphs (98.6%), and larvae (100%) were primarily collected off human hosts.

**Table 1.**
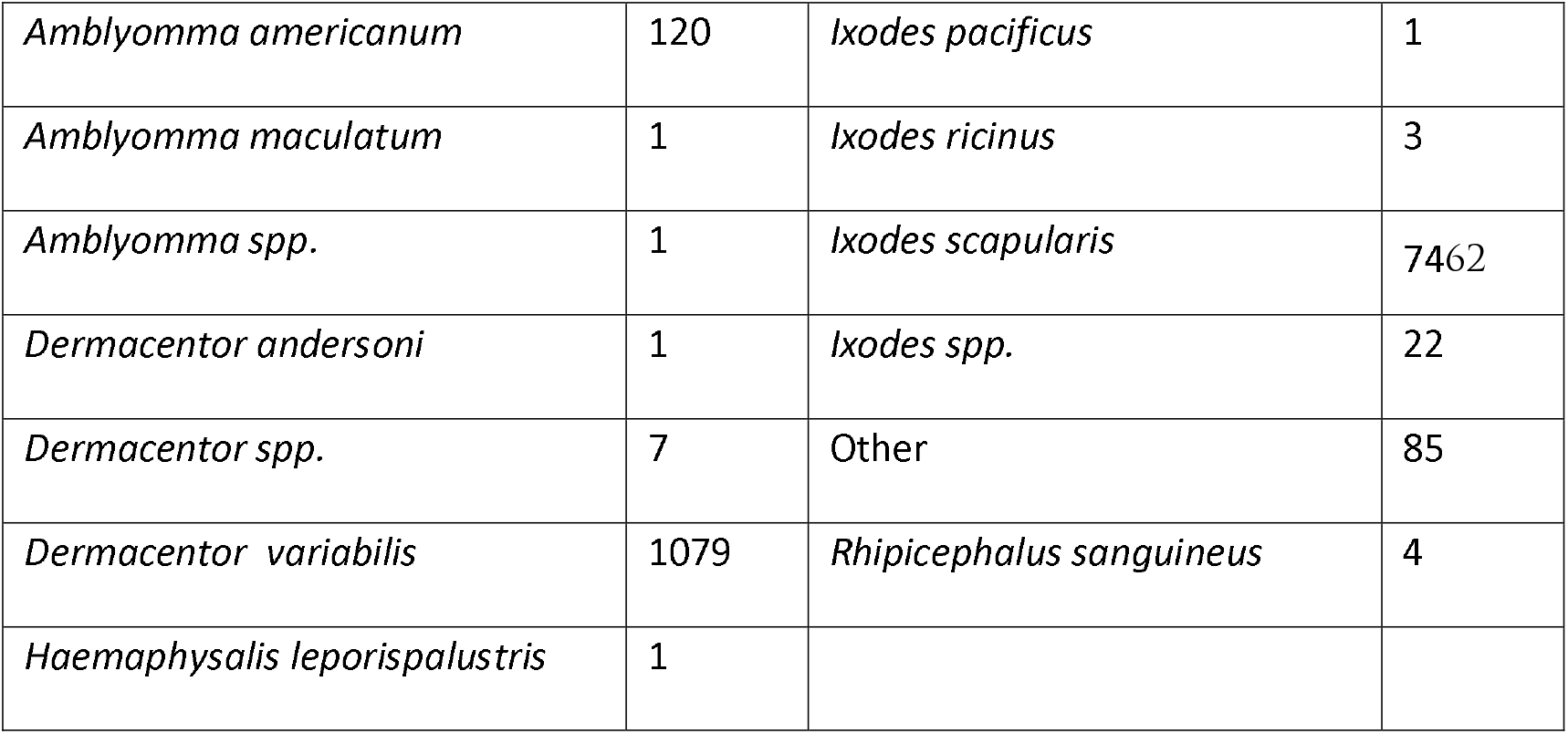
Total Submissions to TickReport from 2015-2017. Submissions of ticks by species as identified by tick species guide.

**Figure 1.** Geographic Coverage of Passive Surveillance. Total number of ticks submitted from every town in Massachusetts from 2015 – 2017.

Of all submitted ticks, 41% (n=3,836) were positive for at least one pathogen and 8.3% (n=773) were positive for two or more pathogens.

The annual prevalence for *B. burgdorferi, B. miyamotoi, B. microti*, and *A. phagocytophilum* are shown in **Table 2**. *B. burgdorferi* and *B. microti* increase in prevalence during this period. *B. miyamotoi* and *A. phagocytophilum* are relatively stable. Our data support previous studies indicating an excess of *B. microti* coinfection with *B. burgdorferi* (χ^2^ =116.35, p-value<0.0001) and *A. phagocytophilum* coinfection with *B. burgdorferi* (χ ^2^ = 12.059, p-value <0.0001) assuming independent transmission in nymphs. There is no significant relationship between *B. miyamotoi* and *B. burgdorferi* in nymphs (χ ^2^ =1.137, p-value=0.286). *Ix. scapularis* adults were submitted every week of the year with peaks in March, May, and November. Nymphs were consistently submitted from April to November. No nymphs were submitted in February. The pathogens follow tick submission patterns with no significant changes in prevalence of any pathogen in either nymphs or adults from month to month.

**Table 2.** Annual prevalence of *B. burgdorferi, B. miyamotoi, B. microti*, and *A. phagocytophilum* in *Ixodes scapularis*. Annual total counts of submitted ticks is given by n. Prevalence, given as p, is calculated from this value for each tick life stage. 95% confidence intervals are given in brackets.

| Year | N | *Borrelia burgdorferi*<br>n (p)<br>[95% CI] | *Borrelia miyamotoi* | *Babesia microti* | *Anaplasma phagocytophilum* |
| --- | --- | --- | --- | --- | --- |
| *Adults* |  |  |  |  |  |
| **2015** | 1543 | 572 (37.1)<br>[34.7, 39.5] | 34 (2.2)<br>[1.5, 3.1] | 88 (5.7)<br>[4.6, 7.0] | 118 (7.6)<br>[6.4, 9.1] |
| **2016** | 1444 | 480 (33.2)<br>[30.8, 35.7] | 30 (2.1)<br>[1.4, 3.0] | 111 (7.7)<br>[6.4, 9.2] | 117 (8.1)<br>[6.7, 9.6] |
| **2017** | 2842 | 1111 (39.1)<br>[37.3, 40.9] | 62 (2.2)<br>[1.7, 2.8] | 230 (8.1)<br>[7.1, 9.2] | 205 (7.2)<br>[6.3, 8.2] |
| *Nymphs* |  |  |  |  |  |
| **2015** | 478 | 91 (19.0)<br>[15.6, 22.8] | 5 (1.0)<br>[0.3, 2.4] | 26 (5.4)<br>[3.6, 7.9] | 24 (5.0)<br>[3.2, 7.4] |
| **2016** | 387 | 78 (20.2)<br>[16.3, 24.5] | 3 (0.8)<br>[0.2, 2.2] | 20 (5.2)<br>[3.2, 7.9] | 15 (3.9)<br>[2.2, 6.3] |
| **2017** | 677 | 162 (23.9)<br>[20.8, 27.3] | 13 (1.9)<br>[1.0, 3.3] | 38 (5.6)<br>[4.0, 7.6] | 27 (4.0)<br>[2.6, 5.7] |

Total tick submissions were 73% adults, 25% nymphs, and 2% larvae. In adult ticks, 44% of submission contained at least one pathogen and 26.6% of nymphs contained at least one pathogen. Polymicrobial infections were found in 8.7% of adults and 5.4% of nymphs. The most common single infection in adults (28.7%) and nymphs (16.7%) was *B. burgdorferi*. The second most common infection status was coinfection with *B. burgdorferi* and *B. microti*: adults (3.9%) and nymphs (3.2%). *A. phagocytophilum* infected ticks included 3.4% of adults and 2.6% of all nymphs. Single infections with *B. microti* and *B. miyamotoi* made up 2.3% and 1.1% of adult ticks and 1.6% and 0.8% of nymph ticks. Three adult ticks were collected that were positive for all four pathogens. Of the 73 larvae submitted, the only positive larva contained *B. miyamotoi*.

### Correspondence of human-biting ticks and case data

The data collected through TickReport on the location and prevalence of tick-borne pathogens are geographically and temporally related to the case report data from the Massachusetts Department of Public Health. To explore the relationship between ticks and cases by geographic location, cases and ticks were aggregated by town and Spearman’s rank (rho) correlation calculated (**Table 3**). Across all data, the tick counts significantly correlate with the reported cases at the town-level. Infected nymphs have the greatest correlation with babesiosis case counts and infected adults have the greatest correlation with anaplasmosis case counts.

**Table 3.** Correlation between Tick Data and Reported Cases. The Spearman’s rank (rho) correlation coefficient is given in the table with its corresponding p-value in parenthesis. The infected nymph and adults refers to the respective pathogens for each disease, *B. microti* infected tick for babesiosis and *A. phagocytophilum* infected ticks for anaplasmosis.

|  | **Babesiosis Cases** | **Anaplasmosis Cases** |
| --- | --- | --- |
| **Adult Submitted** | 0.352 ($p= 3.9e-11$) | 0.404 ($p= 2.8e-14$) |
| **Nymph Submitted** | 0.384 ($p= 4.2e-13$) | 0.360 ($p= 2.2e-11$) |
| **Infected Adults** | 0.303 ($p= 1.8e-8$) | 0.417 ($p= 3.9e-15$) |
| **Infected Nymphs** | 0.371 ($p= 3.2e-12$) | 0.213 ($p= 9.2e-05$) |
| **Total Infected Ticks** | 0.353 ($p= 4.1e-11$) | 0.405 ($p= 3.4e-14$) |

Geographically, the patterns of cases and tick positivity are correlated by infectious agent. While Lyme disease, babesiosis, and anaplasmosis cases are reported throughout the state, *Borrelia miyamotoi* cases are largely restricted to the Cape Cod area. However, all four pathogens have tick submissions from across the state. Comparison of town-level case data (a) and percent positive ticks (b) are shown for Lyme disease (**Figure 2**), babesiosis (**Figure 3**), anaplasmosis (**Figure 4**), and *B. miyamotoi* (**Figure 5**). These figures demonstrate a clear relationship between the geographic distribution of these infectious agents in ticks and in reported disease which visually support the Spearman ranks described in **Table 3**.

**Figure 2.** (a-b) Geographic distribution of Lyme disease cases and *B. burgdorferi* in ticks. (A) Prevalence of Lyme disease cases per 10,000 persons from 2015. (B) Prevalence of *B. burgdorferi* positive ticks per town for towns submitting greater than 5 ticks from 2015 to 2017.

**Figure 3.** (a-b) Geographic distribution of babesiosis cases and *B. microti* in ticks. (A) Prevalence of babesiosis cases per 10,000 persons from 2015 to 2017. (B) Prevalence of *B. microti* positive ticks per town for towns submitting greater than 5 ticks from 2015 to 2017.

**Figure 4.** (a-b) Geographic distribution of anaplasmosis cases and *A. phagocytophilum* in ticks. (A) Prevalence of anbaplasmosis cases per 10,000 persons from 2015 to 2017. (B) Prevalence of *A. phagocytophilum* positive ticks per town for towns submitting greater than 5 ticks from 2015 to 2017.

**Figure 5.** (a-b) Geographic distribution of *B. miyamotoi* disease cases and *B. miyamotoi* in ticks. (A) Prevalence of *B. miyamotoi* cases per 10,000 persons from 2015 to 2017. (B) Prevalence of *B. miyamotoi* positive ticks per town for towns submitting greater than 5 ticks from 2015 to 2017.

### Regression models demonstrate significant link between passive tick surveillance and TBD case reporting

By accounting for the town-level infected adults ticks and the average of positive ticks in each neighboring town, we are able to significantly correlate town-level tick prevalence of *A. phagocytophilum* and reported cases. We observe that a single tick increase in *A*.*p*. positive adult ticks results in an 8.2% increase in cases (IRR=1.082 [1.06-1.11]). Inclusion of spatially lagged averages of the infected adults was further informative, indicating that an average increase in neighboring towns was correlated with a 20% increase in reported cases (IRR=1.20 [1.14-1.27]). Combining these two statistics marginally improved our model (Log-likelihood Ratio χ^2^ = 3.4, p-value=0.066). Based on a map of the residuals of our lagged positive adult model, our model fails to detect the high case load in southwestern Massachusetts. Notably, this region contains some of the only municipalities where we lack any tick submissions.

In summary, positive adult *Ix. scapularis* are spatially correlated with increase in anaplasmosis cases. While the town-only values are significantly correlated, the relationship between tick and human infectivity is more strongly observed with the inclusion of neighboring towns.

The number of submitted nymphs, submitted adults, spatially lagged counts of nymphs, and spatially lagged adult counts had a moderate, though significant, predictive relationship with babesiosis incidence rates. *B. microti* positive nymphs (IRR= 1.75 [1.60, 1.92]) and infected adults (IRR=1.09 [1.07, 1.11]) strongly correlated with babesiosis incidence. Inclusion of infected adults did not improve the infected nymph model (LLR=0.072, p-value=0.832). Incorporating the lagged positive nymph rate slightly improved the nymph model (LLR=7.63, p-value=0.006). In the final model, a town-level increase of 1 positive nymph increased the risk by 57% (43 - 72%) and neighbor-level positive nymphs increase risk by 136% (79-213-%).

In summary, submitted nymph and adult *Ix. scapularis* are correlated with babesiosis case reports on the town level. The total number of *B. microti -*positive nymphs and adults are better predictors of case count than measures of submitted tick density alone. Neighboring towns have similar risk and incorporating a spatially lagged measure of ticks better improves the predictive power of the model.

*A description of the intermediary anaplasmosis and babesiosis models and construction approach is in* ***Supplementary Table 1 and 3***. *The output from all log-likelihood ratio tests is shown in* ***Supplementary Table 2 and 4***.

### Temporal relationship between cases and pathogens

As expected, mid-summer peaks in Lyme disease (**Figure 6a**) and babesiosis (**Figure 6c**) fall between the spring and fall peaks of infected adults and are about a month lagged behind the infected nymph peak. *B. miyamotoi* cases and infected ticks are about 10-fold lower than the other pathogens, making it difficult to draw conclusions about the significance of the possible bi-modal distribution of cases, though tick distribution is as expected (**Figure 6b**). Anaplasmosis displays a unique pattern with an infected nymph peak lagging behind the June peak of cases and a clear bimodal distribution of disease reporting (**Figure 6d**). This finding alongside the output from the counts demonstrate a particular risk for anaplasmosis transmission from adult ticks.

**Figure 6.** (a-d) Temporal Relationship of Infected, Submitted, Blood-fed Adults and Nymphs and Reported Tickborne Diseases. Figures show the annual distribution of submissions for positive-ticks and disease for *B. burgdorferi* in 2015 (a), and averaged monthly from 2015 to 2017 for *B. miyamotoi* (b), *B. microti* (c), and *A. phagocytophilum* (d). Nymphs are given by the dotted yellow line, adults in the solid yellow line, and case reports in dotted blue.

## Discussion

The TickReport passive surveillance database serves an essential function for measuring and reporting public risk of tick-borne disease. A systematic review of various flagging studies of *Ix. scapularis* determined the average prevalence of *Borrelia miyamotoi* in Massachusetts to be 2.5%, *Babesia microti* to be 5.3% and *Borrelia burgdorferi* was found to be between 5-16% (Nelder et al. 2016). A recent report relying on passive surveillance found *A. phagocytophilum* prevalence in Massachusetts to be 9.2%, *B. microti* at 2.6%, *B. burgdorferi* at 26.9%, and *B. miyamotoi* at 1.7% (Nieto et al. 2018).

A publication on TickReport data from Massachusetts from 2006-2012 reported 63% of human-biting ticks carried at least one pathogen and 8% carried two pathogens. In this analysis, 41% of ticks were infected with at least one pathogen, 8.3% were infected with more than 1 pathogen, and 1.7% were infected with more than 2 pathogens. The prevalence of *B. burgdorferi, A. phagocytophilum*, and *B. microti* in this population was 29.6%, 4.6%, and 1.8%, respectively (Xu et al. 2016). In ticks submitted to TickReport from 2006–2012, 29.6% *Ix. scapularis* were infected with *B. burgdorferi*, 4.6% with *A. phagocytophilum*, and 1.8% with B. *microti* (Xu, unpublished). For 2015-2018, 30.39%-36.19% Ixodes scapularis were infected with *B. burgdorferi*, 6.98%-7.32% with *A. phagocytophilum*, and 5.65%-8.18% with *B. microti*. This indicates a significant increase in prevalence of all three pathogens. Data on *B. miyamotoi* for the prior period are not available.

The observation of a single larva positive for *B. miyamotoi* is a useful contribution to the study of transovarial transmission in *Ix. scapularis*. This supports work presented on the high transovarial rate of 90.9% recently published by Han et al. (Han 2019). *B. burgdorferi* and *A. phagocytophilum* are not known to be transmitted transovarially (Dunning Hotopp, 2006) and no larvae in the present study were found infected with either of these pathogens.

The negative binomial regression count models of the relationship between tick submissions and case reports indicates these data are highly valuable for public health surveillance. The town-level spatial resolution is achieved due to the nature of this passive reporting dataset and gives a more accurate view of human risk, given the proximity of the submitted ticks and the human host. Previous studies have found a difference in *B. burgdorferi* infection prevalence within *Ixodes* ticks removed from humans compared to those collected by flagging from the same region (Waindok et al., 2017). Recent work has demonstrated a significant correlation between the number of submitted *Ixodes scapularis* ticks and the reporting of Lyme disease cases (Gasmi et al. 2019). The context of this work was the expanding edge of the tick habitat whereas this work addresses already endemic regions. In Connecticut, *B. burgdorferi* positive ticks and tick density were highly correlated with disease reporting (Stafford 1998). However, historical flagging studies have failed to associate infection prevalence in ticks with reported cases (Daniels et al., 1998). This analysis was limited to the regional scale. Our models identify the added predictive power of pathogen load information on these submitted ticks across two additional pathogens (i.e. we correct for the number of ticks submitted – which is significant here as well- and we still find contribution from the number of positive ticks – confirmed by log-likelihood test). Further, Dunn et al. found that tick burden alone was not predictive of the expansion of *Borrelia burgdorferi, Babesia microti*, and *Anaplasma phagocytophilum* (Dunn et al. 2013).

The absence of a second peak Lyme disease and babesiosis in the fall could indicate a shorter attachment time is required for *A. phagocytophilum* transmission. There may be an additional effect from the lack of physician awareness about continuing risk for tick-borne diseases following the summer months. However, this was difficult to evaluate with these data as reported cases of Lyme disease were only available for the year 2015, while anaplasmosis and babesiosis cases were averaged using reports from 2015 to 2017.

A review of transmission studies found that in experiments with a single *B. burgdorferi*-infected tick, 0% of experimental rodent hosts were infected after 24 hours of attachment, but that 53% of hosts were infected by 63-67 hours (Eisen 2018). In a single study of *B. miyamotoi* by a single infected nymph, 10% of rodents were infected within 24 hours and 63% after 72 hours. *A. phagocytophilum* was successfully transmitted to 67% of rodents by 36 hours and 100% by 72 hours. *B. microti* transmission studies rely on one study from 1987 where it was found that 71% of rodents were infected after 54 hours of feeding.

The presence of a second case peak of anaplasmosis in the fall draws interest to the potential role of adult ticks in transmission of *A. phagocytophilum*. In the anaplasmosis models, pathogen-positive adults more significantly contributed to our case predictions whereas the babesiosis model is significantly driven by nymph data. These data contradict the presumption that nymphs typically feed longer, allowing for more transmission time. In this dataset, adults and nymphs had similar engorgement rates. Therefore, the understanding of nymphs as the “principal vector” of tick-borne disease may be incomplete in the case of *A. phagocytophilum*.

Previous work in New Hampshire using sampling sites has identified a significant correlation between entomologic risk index (total number of ticks x proportion of ticks infected) and incidence of human Lyme disease (Walk, Stull, & Rich 2009). Globally, this method of public health surveillance has gained attention due to its spatial scale and importance for risk mapping (Gasmi et al., 2019; Garcia-Marti et al., 2018). Passive surveillance data on *Ix. scapularis* ticks provide essential information about disease risk. Where tick questing behavior has been shown to significantly impact Lyme disease risk (Arsnoe et al. 2019) access to geotagged data about ticks that are directly interacting with human hosts is essential for public health surveillance.

Here we show the value of this system for monitoring true public health risk beyond flagged nymphs. As described by Walk et al. in 2009 (Walk, Stull, & Rich 2009), adult ticks should be considered as relevant, if not more so given our models, as nymphs for assessing public health risk. When we report combined nymph and adult prevalence, we are aware that adult infection rate is about twice of nymph infection rate (Barbour & Fish 1993) and therefore may reflect an increase in adult tick submissions. However, when pertinent data are split by tick stage, our data demonstrate that significant prevalence changes are due to an increase across tick stages and not merely an increase in adults.

An important caveat in comparing the reported tick location and the public health data is that tick data are linked to the reported tick bite location whereas public health data are reported based on the individual’s reported town of residence. We previously showed that many instances of *Borrelia burgdorferi* positive TickReport submissions from the West coast (California, Oregon, and Washington) involved bite of non-endemic ticks acquired while human subjects were travelling in the Northeast of upper Midwest where *Ix. scapularis* ticks are endemic (Xu et al, 2019). In the present dataset, 72% of ticks were reported in the same town as the submitter’s home address. Since the two locations are preserved for each TickReport record, the location disparity can be recorded and measured directly where such provisions are not possible in case reports.

TickReport provides a unique surveillance tool for monitoring adults, nymphs, and to some extent larvae that have confirmed contact with humans. The accounting of risk components among the human biting ticks gives a unique picture of human disease risk not possible with traditional flagging methods. Moreover, because the data are generated through the fee-for-service, this surveillance approach is sustainably crowd-funded unlike previous efforts relying on ephemeral private grant support (Nieto et al. 2018).

## Data Availability

Passive surveillance data are freely available at www.tickreport.com/stats.

